# Multi-centre validation of MELD-graph for automated focal cortical dysplasia detection using 7T compared to paired 3T MRI

**DOI:** 10.64898/2026.10.08.26365027

**Authors:** Cornelius Kronlage, Mathilde Ripart, Rory J Piper, Chiara Casella, Krzysztof Klodowski, Tobias Bauer, Maria H Eriksson, Philippa Bridgen, Martin M Tisdall, Simon R Bouschery, Annalena Lange, Eberhard D Pracht, Tony Stöcker, Jon O Cleary, Felice D’Arco, Torsten Baldeweg, John S Duncan, Rainer Surges, Theodor Rüber, Christopher T Rodgers, Thomas E Cope, David W Carmichael, Jonathan O’Muircheartaigh, Sophie Adler, Konrad Wagstyl

**Author notes:** joint last authorship (Sophie Adler, Konrad Wagstyl).

## Abstract

**Background:** Software tools can support MRI diagnosis of focal cortical dysplasias (FCDs) – subtle, surgically-treatable lesions that underlie drug-resistant focal epilepsy (DRFE). MELD-graph, a widely used open-source tool combining surface-based morphometry with a convolutional neural network, was developed using 1.5T and 3T MRI data. 7T MRI is increasingly used for lesion detection in DRFE. This study investigated whether MELD-graph is robust to domain shift at 7T.

**Methods:** We collated paired 3T and 7T T1-weighted MRI, and FLAIR where available, from 23 patients with FCD (6 histologically-verified, 17 identified radiologically) and 39 healthy controls at four epilepsy centres. Manual lesion labels were used as ground truth. MELD-graph was run on 3T images, on 7T images with the default pipeline, and on 7T images with adaptations for 7T MRI (SPM bias-field correction, SynthStrip skull-stripping, sub-millimetric FreeSurfer reconstruction), with and without ComBat harmonisation. FCD detection performance and surface morphometry features were compared.

**Results:** Sensitivity for FCD detection was 65% (15/23) at 3T, 48% (11/23) for 7T default and 52% (12/23) for 7T adapted, with Combat harmonisation. Among radiologically 3T-negative patients, 2/8 were detected at 3T and 3/8 at 7T; and among histologically confirmed FCD, 3/6 at 3T and 1/6 at 7T. Specificity was significantly lower at 7T (default 39%, adapted 45%) than 3T (76%). False-positives predominantly occurred in the basal temporal lobes. ComBat harmonisation improved specificity in all conditions and largely compensated for field-strength dependent differences in surface morphometry features. Adding FLAIR at 7T increased false-positive predictions.

**Conclusion:** Sensitivity is comparable but specificity of MELD-graph is lower at 7T compared to 3T. Adaptations for 7T MRI can reduce segmentation errors but do not significantly improve FCD detection, while harmonisation improves performance. The adapted pipeline is released open-source. 7T MELD-graph outputs may inform radiologist review in challenging cases but require cautious interpretation.

## Introduction

Epilepsy surgery can be a curative treatment in drug-resistant focal epilepsy (DRFE). Focal cortical dysplasias (FCDs) are among the most common resected epileptogenic lesions [Blumcke et al., 2017], with favourable postoperative seizure freedom rates of 65-70% [Fauser et al., 2015; Lamberink et al., 2020]. There are different histopathological subtypes, of which ILAE type II FCD is the most prevalent [Najm et al., 2022].

MRI is essential in the pre-surgical diagnostic workup for detecting and locating putative lesions. Radiologically, FCD II are typically characterised by abnormal cortical thickness, blurring of the grey-white matter boundary, changes in T2/FLAIR signal intensity, and the transmantle sign. Other subtypes, such as FCD I, are less conspicuous [Isler et al., 2017]. In general, FCDs can be very subtle and escape radiological diagnosis [Walger et al., 2025]. Among MRI-negative patients who undergo resections on the grounds of other investigations such as intracranial EEG, histopathology most frequently reveals FCD [Bien et al., 2009; Wang et al., 2013]. Improving FCD detection on MRI could avoid more invasive investigations, improve timely access to epilepsy surgery and lead to better outcomes for people with DRFE [Téllez-Zenteno et al., 2010].

One approach is optimising analysis of MRI images. Increasing reader experience and repeated reviews have been shown to improve diagnostic yield [Walger et al., 2024; Wehner et al., 2021]. Furthermore, computational approaches for supporting diagnosis have been developed, ranging from statistical voxel- or surface-based morphometry methods [Wagner et al., 2011] to deep-learning algorithms [Gill et al., 2021; Kersting et al., 2025; Ripart et al., 2025]. These have various strengths and limitations in terms of performance, time required for processing and review of the outputs, and interpretability of the predictions. Even though none of these tools have undergone medical device regulation, some are routinely used at epilepsy surgery centres to support radiologist review of brain MRI in challenging cases [Adler et al., 2025], a practice that is supported by expert consensus recommendations [Bernasconi et al., 2019].

Another approach focuses on image acquisition. 7T MRI is increasingly entering clinical practice for lesion detection in DRFE [Opheim et al., 2021], offering higher spatial resolution and signal-to-noise ratio, at the cost of stronger artefacts arising from radiofrequency field inhomogeneity [Ladd et al., 2018]. 7T MRI is well documented to enable more sensitive radiological diagnoses. Recent studies in DRFE have reported diagnostic gains of 23-31% [Klodowski et al., 2025; van Lanen et al., 2021; Vecchiato et al., 2025], with FCD the most common histopathological diagnosis.

Applying computational post-processing and FCD detection tools to 7T MRI requires careful validation. All such tools are statistical models fitted to reference data. An algorithm developed with, for instance, 3T MRI data does not necessarily generalise to 7T MRI with its different characteristics – a phenomenon known as ‘domain shift’ [Hoffmann, 2025]. This can be addressed by development of 7T MRI-specific versions, which is feasible for statistical approaches that require only a small amount of healthy control data [Chen et al., 2021; Wang et al., 2020]. However, the performance of modern deep learning algorithms depends on the size, quality and diversity of the labeled dataset used for training. Whereas multi-centre 1.5T/3T datasets can comprise hundreds of FCDs [Gill et al., 2021; Ripart et al., 2025], 7T cohorts are still an order of magnitude smaller [Burkett et al., 2026; Hangel et al., 2024; van Lanen et al., 2021; van Lanen et al., 2025]. Therefore, at this point in time, applying deep learning algorithms trained on large and diverse 3T datasets could help leverage and transfer performance advantages to 7T MRI.

MELD-graph is a widely used open-source research software for automated FCD detection [Ripart et al., 2025] that has performed favourably in comparison to other tools in independent validation studies [Bücheler et al., 2026; Kaas et al., 2026]. It was developed using 1.5T and 3T MRI data of 703 FCD patients and 482 controls. MELD-graph uses FreeSurfer [Fischl, 2012] for extraction of interpretable cortical morphometry features, which undergo statistical normalisation and harmonisation for site-specific effects before predictions are generated by a graph-convolutional neural network. It is unclear to what degree MELD-graph might be susceptible to domain shift. We found that a tool for hippocampal sclerosis detection with similar overall design performed robustly at 7T [Kronlage et al., 2026].

In a recent study, a customised version of MELD-graph with adaptations tailored for 7T MRI was tested on a dataset of 87 DRFE patients with 7T T1-weighted and FLAIR MRI acquired at a single site. Overall higher diagnostic performance at 7T was described, although segmentation errors, which may be more common at 7T and likely to cause false positive predictions, were manually excluded [Lenge et al., 2026].

In this study, we systematically evaluated the performance of MELD-graph for FCD detection using 7T MRI from four tertiary care epilepsy centers (Figure 1). For comparison, 3T MRI data of the same patients were analysed. We investigated to what degree a preprocessing and segmentation pipeline adapted for 7T MRI, ComBat harmonisation of surface morphometry features and inclusion of FLAIR had an effect on downstream robustness of FCD detection at 7T. We make the adapted processing pipeline available open-source as part of the MELD-graph package.

**Figure 1:**
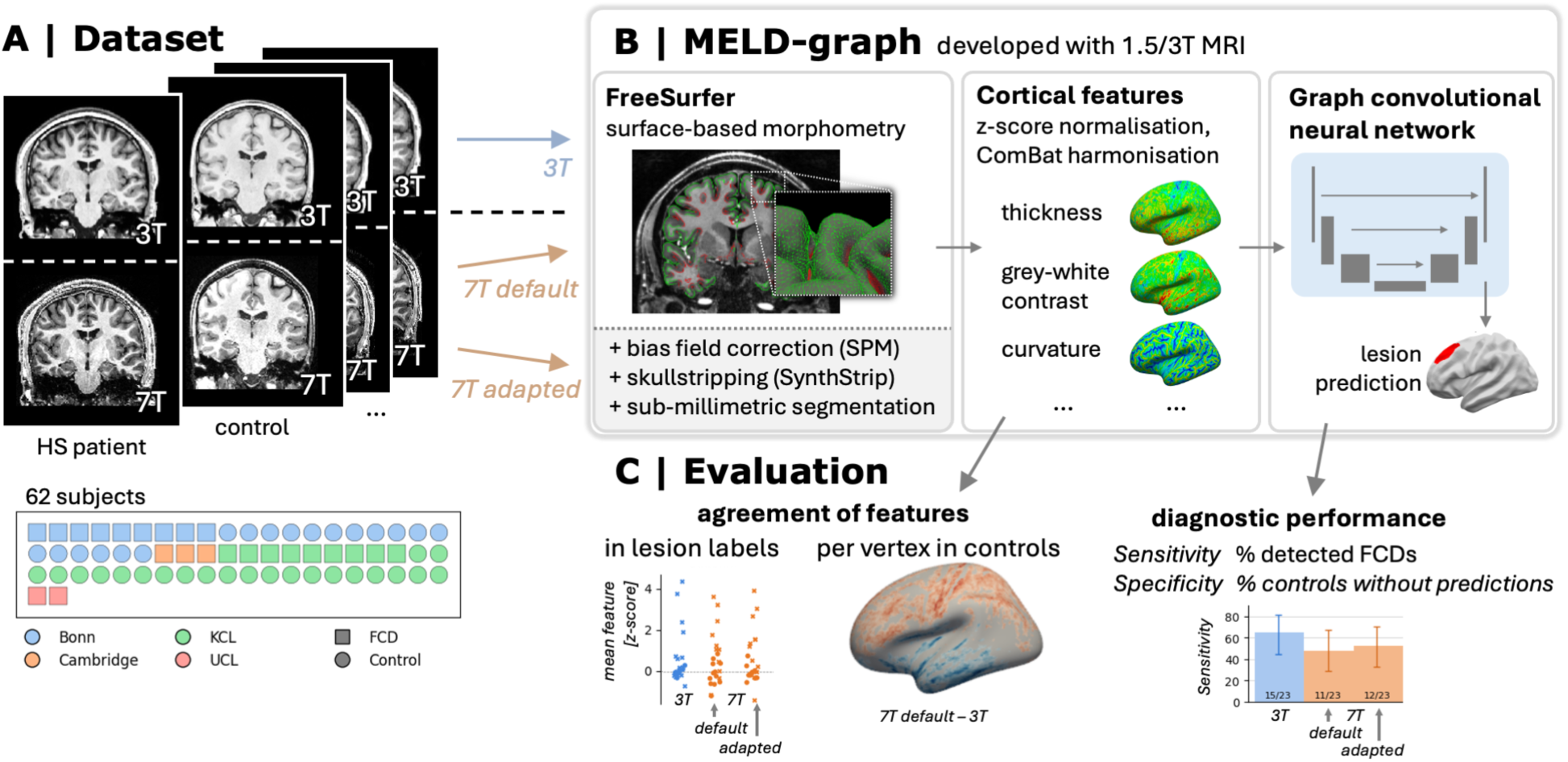
Study overview. (A) The dataset comprised paired 3T and 7T MRI T1-weighted (and FLAIR) images of 23 patients with focal cortical dysplasia (FCD), with manual lesion labels, and 39 healthy controls. (B) The open-source software MELD-graph was applied independently to 3T images, 7T images (‘7T default’) and 7T images with adapted preprocessing and segmentation steps (‘7T adapted’). (C) Evaluation included assessment of diagnostic performance – sensitivity for detection of FCDs and specificity in healthy controls – and agreement of surface morphological features between field strengths and processing conditions.

## Methods

### Dataset

Paired 7T and 3T MRI and clinical data were collected from four different epilepsy centres, where DRFE patients and healthy controls had undergone 7T MRI for clinical or research purposes. Ethics committee approvals for each site were: Cambridge: UK National Research Ethics Service (23/WM/0008). Bonn: Ethics Committee at the Medical Faculty of the University of Bonn (083/20, 2025-55-BO). KCL: UK Health Research Authority (IRAS 243811), UK National Research Ethics Office (ref. 18/LO/1766). UCL: Bradford Leeds Research Ethics Committee (REC ref: 22/YH/0171), King’s College London Research Ethics Committee (Development of Novel Magnetic Resonance Techniques Using Healthy Volunteers Study, KCL REMAS ref: 8700). The analysis was approved by the UK Health Research Authority (IRAS 301863).

Individual cohorts and data acquisition have been described previously: University College London, UK (UCL); King’s College London, UK (KCL) [Dokumacı et al., 2023; Vecchiato et al., 2025]; University of Cambridge, UK [Klodowski et al., 2025]; and University Hospital Bonn, Germany [Faber et al., 2026], as well as the collated dataset [Kronlage et al., 2026].

Patients with focal cortical dysplasia (FCD) and healthy controls were selected for this study. FCD patients were eligible if they met the following criteria: diagnosis of DRFE and radiologically identified FCD on either 3T or 7T MRI, with multidisciplinary team consensus, or histopathologically verified FCD. Healthy controls were required to have no neurological diagnosis and to have undergone 7T MRI using the same imaging protocol as patients. Across both groups, subjects were required to have both 7T and 3T T1-weighted MRI images available. We excluded subjects with uninterpretable images, e.g., due to movement artefacts, based on manual quality control.

Lesion labels for FCDs were manually drawn by neurologists and/or neuroradiologists (using ITK-SNAP version 4.4.0) on the available T1-weighted or FLAIR image that provided the clearest visualisation of the lesion, using other clinical imaging and documentation (such as radiological reports, multidisciplinary meeting notes and postoperative MRI) for reference.

Additionally, the following demographic and clinical variables were collected and analysed: age, sex, radiological findings (visually positive/negative with respect to FCD) on 3T MRI and 7T MRI, whether surgery was performed, histology, and favourable outcome (defined as Engel or ILAE class I at least one year after surgery).

### Ultra-high field MRI adaptations of MELD-graph

We adapted MELD-graph [Ripart et al., 2025] to perform optional preprocessing steps at the segmentation stage for ultra-high field 7T MRI data: (1) In case of uniform MP2RAGE with background noise as inputs, estimation of de-noised, ‘robust’ images using the protocol described in [O’Brien et al., 2014]. (2) Rescaling of images to positive values, because MP2RAGE can contain negative values that disrupt later processing steps. (3) Bias-field correction using the unified segmentation method of SPM (version 25.01.02) [Ashburner and Friston, 2005]. (4) Skull-stripping with SynthStrip, using the ‘--no-csf’ model to include the minimum amount of dura in the brain mask. (5) Surface-based morphometry using freesurfer (version 7.2.0), with volume segmentation at the native, sub-millimetric resolution (‘-hires’ flag) [Fischl, 2012].

### Processing

For each subject, the MELD-graph segmentation and feature extraction was performed in three separate runs: with the 3T T1-weighted image as input (referred to as ‘3T’), with the 7T T1-weighted image without preprocessing or changes to the pipeline (‘7T default’), and with the 7T image using the adaptations described above (‘7T adapted’) (Figure 1). For each condition, predictions were then generated both with and without ComBat harmonisation for site-specific biases, yielding a total of six predictions per subject.

Additionally, we obtained predictions using both T1-weighted and FLAIR inputs, acquired at 3T (‘3T FLAIR’) – or 7T and processed with the proposed adaptations (‘7T FLAIR’).

### Evaluation of predictions

Manual lesion masks were defined in the voxel space of different 3T or 7T images in each patient case. For evaluation, they had to be aligned to the T1-weighted 3T/7T volumes used as primary inputs for MELD-graph, and thereby to the MELD-graph voxel-wise predictions. We skullstripped each pair of source and target images using SynthStrip [Hoopes et al., 2022]), estimated an affine registration using EasyReg [Iglesias, 2023] and applied this to resample the lesion mask volume with nearest neighbour interpolation.

MELD-graph predicted clusters were evaluated as true- or false-positive using any voxel overlap with a lesion mask as criterion. We calculated the following subject-wise performance metrics:

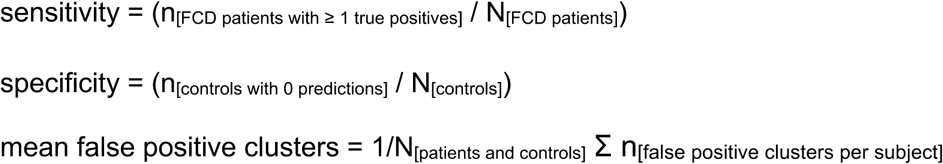

95% confidence intervals were computed as Wilson binomial proportion intervals (sensitivity, specificity) or by bootstrapping (random re-sampling with replacement, 10,000 iterations).

Differences between paired conditions were assessed with McNemar tests (for sensitivity and specificity) or Wilcoxon-signed-rank tests (for mean false positive clusters), with Benjamini-Hochberg correction of p-values for a false-discovery rate (FDR) of 0.05.

To reflect how predictions obtained at both field strengths could be interpreted clinically, we evaluated a combined ‘3T + 7T adapted’ condition. For the combination, an FCD was considered ‘detected’ if there was a prediction at either field strength that overlapped with the ground truth lesion mask. False-positive predictions at both field strengths were summed.

We assessed size (measured in cluster vertices) and confidence scores of predictions, as well as DICE scores between manual lesion masks and MELD-Graph predictions (measured by number of vertices on the template ‘white’ surface), and compared distributions between conditions computing a Kruskal-Wallis-test.

### Visualisation of example cases with predictions

To allow for a side-by-side comparison of visual features and predictions, 3T images and MELD-graph predictions were co-registered and resampled to corresponding 7T images (by first skullstripping using SynthStrip [Hoopes et al., 2022] to improve robustness for MP2RAGE images, and then affine coregistration using FreeSurfer mri_coreg). Slices centered on the FCD lesion labels or predictions were plotted.

### Spatial distribution of lesions and false positive predictions

Ground truth lesion masks and false-positive MELD-graph predictions from different conditions were sampled to a template cortical surface mesh (FreeSurfer fsaverage_sym). To visualise the spatial distribution of lesions and predictions in different conditions (3T, 7T default, 7T adapted), we summed their number at each vertex over both hemispheres and plotted this sum overlaid on surface renders. To visualise the whole data range, we chose a power-law gamma colorscale (value_colorscale_ = count^γ^ with γ=0.5).

### Surface morphometry features

We explored to what degree differences of extracted surface morphometry features between field strengths might have underpinned differences in prediction performance. We used the features pre-processed by ComBat harmonisation (if applicable, in the ComBat conditions) as well as intra- and inter-subject z-score normalisation, termed ‘normalised’ features in the MELD-graph pipeline. Values were mapped to the symmetrical fsaverage_sym template, which allowed comparisons of lesional areas across field strengths and conditions or homotopic cortical areas over the whole cortex across healthy controls. We conducted two analyses: Firstly, we compared average feature values in the manual FCD labels across conditions and assessed the differences in paired distributions using Friedman tests. Secondly, we assessed per-vertex differences of features across all healthy controls between the 7T default vs. 3T, and the 7T adapted vs. 3T conditions by computing Wilcoxon-signed-rank tests with Benjamini-Hochberg FDR-correction of p-values; we show surface overlay plots of the median difference only for vertices with p_FDR_<0.05.

### Data and code availability

We are unable to make the MRI dataset publicly available due to data protection restrictions. A fork of MELD-graph (version 2.2.7) with optional adaptations for ultra-high field MRI segmentation is available at https://github.com/ckronlage/meld_graph/tree/v2.2.7_uhf along with usage instructions. Scripts for the analyses presented in this manuscript can be accessed at https://github.com/ckronlage/meldgraph_uhf_pub.

## Results

### Cohort and dataset

The cohort with paired 7T and 3T data comprised 62 subjects. A breakdown of demographic and clinical variables is shown in Table 1. There were 23 patients with FCD, of which 6 were verified by histopathology and 17 identified radiologically. Mean age was 19.6 years (median 15 years, range 8-43). There were 37 male and 25 female participants. There were no statistically significant differences in age and sex between groups (Chi-square test, Mann-Whitney-U test).

**Table 1:**
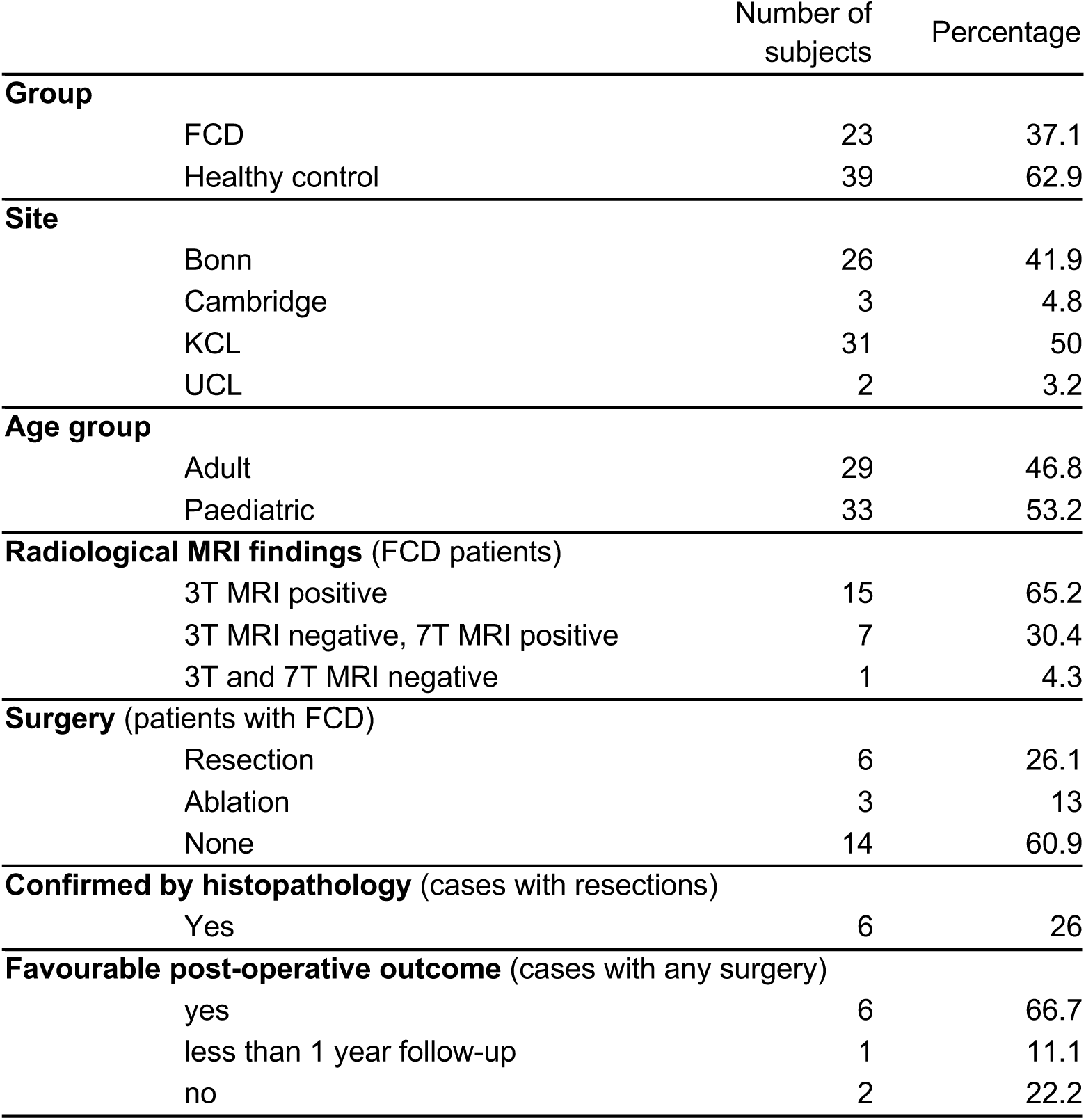
Multi-centre paired 7T and 3T MRI focal epilepsy cohort, demographic and basic clinical data.

|  | Number of subjects | Percentage |
| --- | --- | --- |
| **Group** |  |  |
| FCD | 23 | 37.1 |
| Healthy control | 39 | 62.9 |
| **Site** |  |  |
| Bonn | 26 | 41.9 |
| Cambridge | 3 | 4.8 |
| KCL | 31 | 50 |
| UCL | 2 | 3.2 |
| **Age group** |  |  |
| Adult | 29 | 46.8 |
| Paediatric | 33 | 53.2 |
| **Radiological MRI findings (FCD patients)** |  |  |
| 3T MRI positive | 15 | 65.2 |
| 3T MRI negative, 7T MRI positive | 7 | 30.4 |
| 3T and 7T MRI negative | 1 | 4.3 |
| **Surgery (patients with FCD)** |  |  |
| Resection | 6 | 26.1 |
| Ablation | 3 | 13 |
| None | 14 | 60.9 |
| **Confirmed by histopathology (cases with resections)** |  |  |
| Yes | 6 | 26 |
| **Favourable post-operative outcome (cases with any surgery)** |  |  |
| yes | 6 | 66.7 |
| less than 1 year follow-up | 1 | 11.1 |
| no | 2 | 22.2 |

The 7T MRI sequence characteristics have been previously described for this collated cohort [Kronlage et al., 2026] and detailed site specific acquisition parameters are published [Dokumacı et al., 2023; Faber et al., 2026; Klodowski et al., 2025; Vecchiato et al., 2025]. Briefly, 3D T1-weighted images had voxel sizes between 0.65 and 0.8 mm (median 0.65 mm), included MPRAGE (Bonn, n=26) and MP2RAGE (other sites, n=36) sequences, were acquired with parallel transmit (pTx) universal pulses (UP) for n=26 and in circularly polarised (CP) mode for n=36 subjects. FLAIR at both 7T and 3T was available for 48 subjects, 3T FLAIR in-plane resolution was 0.5-1.0 mm (median 1.0 mm); 7T FLAIR was acquired using UP in 45 and CP in 3 subjects, in-plane resolution was 0.6-0.8 mm (median 0.6 mm).

### MELD-graph FCD detection performance

Using T1-weighted images as inputs, Freesurfer reconstruction failed in 1/62 subjects for 7T data with the default MELD-graph pipeline, in contrast to none using the adapted processing or 3T inputs.

With ComBat harmonisation, MELD-graph detected 65% (15/23) of FCDs at 3T vs. 48% (11/23) in the ‘7T default’ and 52% (12/23) in the ‘7T adapted’ condition. Of the radiologically 3T MRI-negative patients, MELD-graph detected 25% (2/8) at 3T and 38% (3/8) at 7T (default and adapted). Among histopathologically verified FCDs, sensitivity was 3/6 at 3T and 1/6 for 7T default and adapted.

In healthy controls, specificity was 76% (29/38) at 3T and significantly lower, 39% (15/38) for ‘7T default’ (McNemar FDR-corrected p=0.008) and 45% (17/38) for 7T adapted (p=0.019). Similarly, the mean number of false-positive predictions per subject across patients and controls was 0.2 at 3T vs. 0.6 for 7T default (Wilcoxon signed-rank test p < 0.001) and 0.6 for 7T adapted (p < 0.001). ComBat harmonisation improved specificity at 7T and decreased the number of false-positive predictions in all conditions (p < 0.05). Performance of the 7T adapted pipeline did not differ significantly from the 7T default pipeline. Quantitative performance metrics are shown in Figure 2. True-positive or false-positive prediction clusters did not significantly differ in size, dice score (in relation to the lesion label) or confidence score (Supplementary Figure 1).

**Figure 2:**
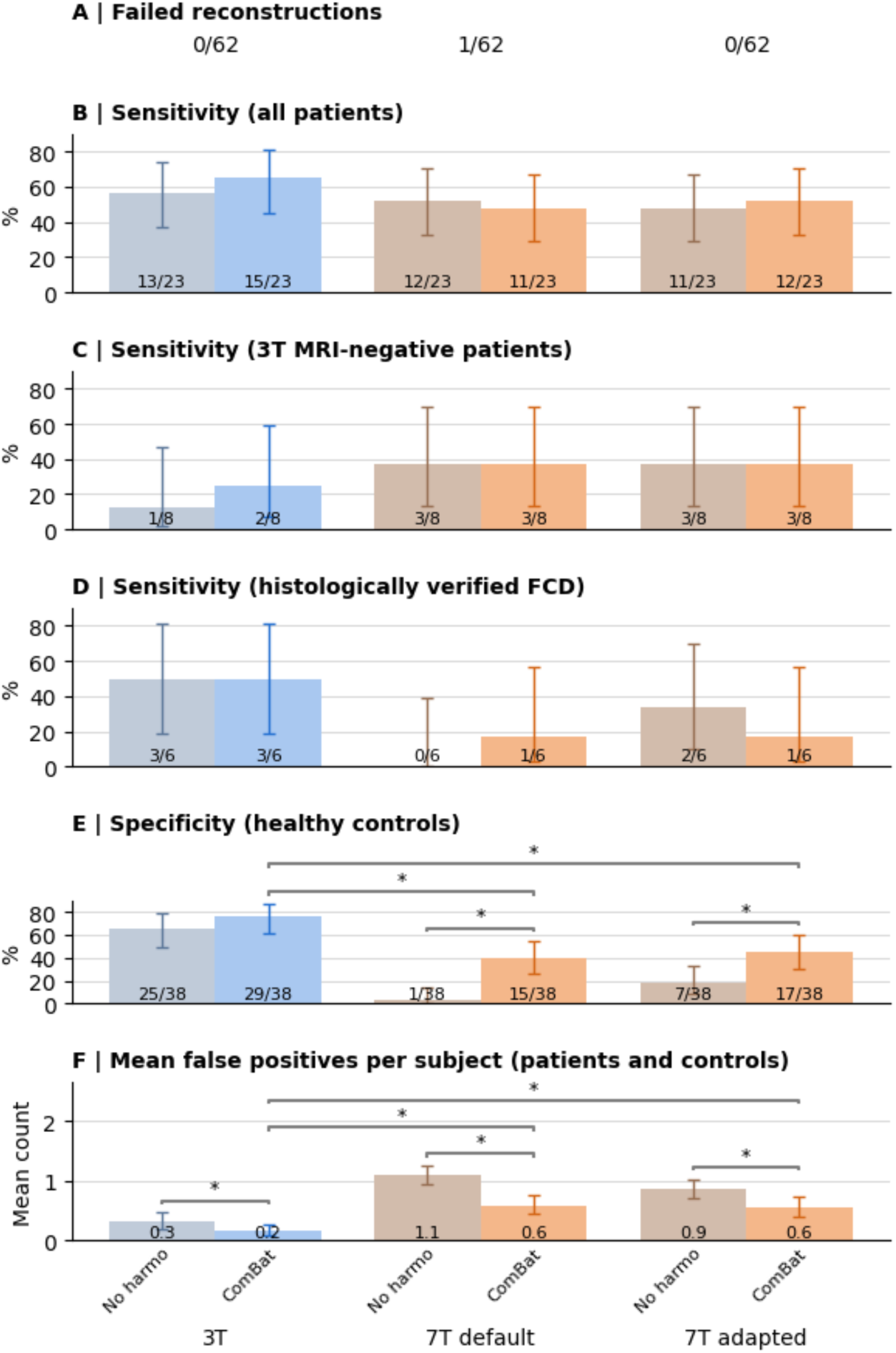
FCD detection performance of MELD-graph using T1-weighted images acquired at 3T (left), 7T processed with the default pipeline (middle) and 7T with adapted preprocessing (right). (A) Fraction of failed Freesurfer reconstructions in each condition; further metrics below are only for n=61 subjects with successful reconstructions under all three conditions. (B) Sensitivity in all FCD patients. (C) Sensitivity in subgroups of radiologically 3T MRI-negative patients and (D) histopathologically confirmed FCD cases. (E) Specificity (no predictions in healthy controls). (F) Mean number of false positive predictions in patients and controls. In each condition, results are shown without (darker shade) and with (lighter shade) ComBat harmonisation to correct site batch effects. Error bars show 95% confidence intervals. Statistical tests were computed for comparisons between the three field strength/processing conditions ‘3T’, ‘7T default’, ‘7T adapted’ with ComBat harmonisation, and between ‘No harmo’ and ‘ComBat’ within each condition. *statistically significant differences (p < 0.05 with Benjamini-Hochberg false discovery rate correction)

The quantitative performance of a combination of 3T and 7T adapted MELD-graph predictions is shown in Supplementary Figure 2: 70% (16/23) FCDs were detected either at 3T or 7T (i.e., one 3T-negative patient was detected at 7T), though specificity decreased to 36% (14/39) and the combined number of false-positives (0.7) was higher.

Detection performance when using 3T FLAIR in addition to 3T T1-weighted images was not statistically different for the subgroup of subjects with 3T FLAIR available. With 7T FLAIR images as additional inputs, specificity was lower than when using 7T T1-weighted images only (Supplementary Figure 3).

Examples of MELD-graph predictions in individual cases are shown in Figure 3, along with prediction confidence scores and FreeSurfer surface overlay plots highlighting segmentation errors for predictions in the temporal lobes. Further examples of false-positive predictions in healthy controls are shown in Supplementary Figure 4.

**Figure 3:**
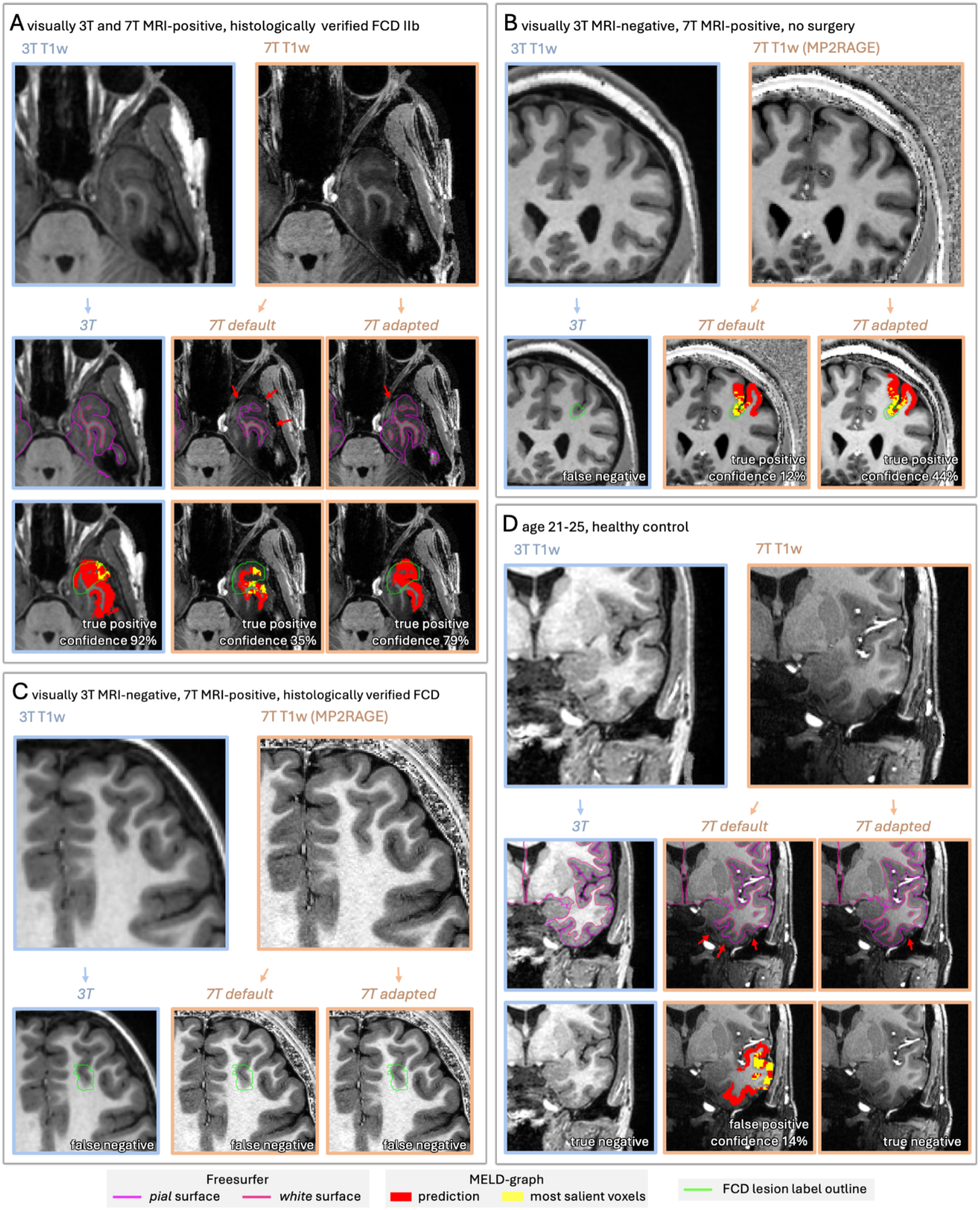
Case examples of MELD-graph predictions, all using ComBat harmonisation. (A) Patient age 11-15 years, visually 3T and 7T MRI positive and histopathologically verified FCD IIb. (B) Patient age 26-30 years, visually 3T MRI-negative, identification of an FCD in the left superior frontal sulcus on 7T MRI, no surgery. (C) Patient age 11-15, visually 3T MRI-negative, identification of an FCD in the left frontal lobe on 7T MRI, histopathologically verified. (D) Healthy control age 21-25 years. For each case, input images without overlays and input images with overlaid lesion label outlines (green outline) and MELD-graph predictions (red/yellow clusters) are shown. For cases A and D, additional images with overlaid Freesurfer surfaces with red arrows showing segmentation errors are displayed. See Supplementary Figure 4 for more examples of false positives in healthy controls.

### Spatial distribution of false-positive predictions

FCD lesions in this cohort were mostly located in the temporal and frontal lobes (Figure 4A). Without harmonisation of features, false-positive MELD-graph predictions in the 7T default and 7T adapted conditions were predominantly located in the lower temporal lobes and at the temporal pole. With ComBat harmonisation, there were overall less false positives, and the temporal predominance was less pronounced (Figure 4B).

**Figure 4:**
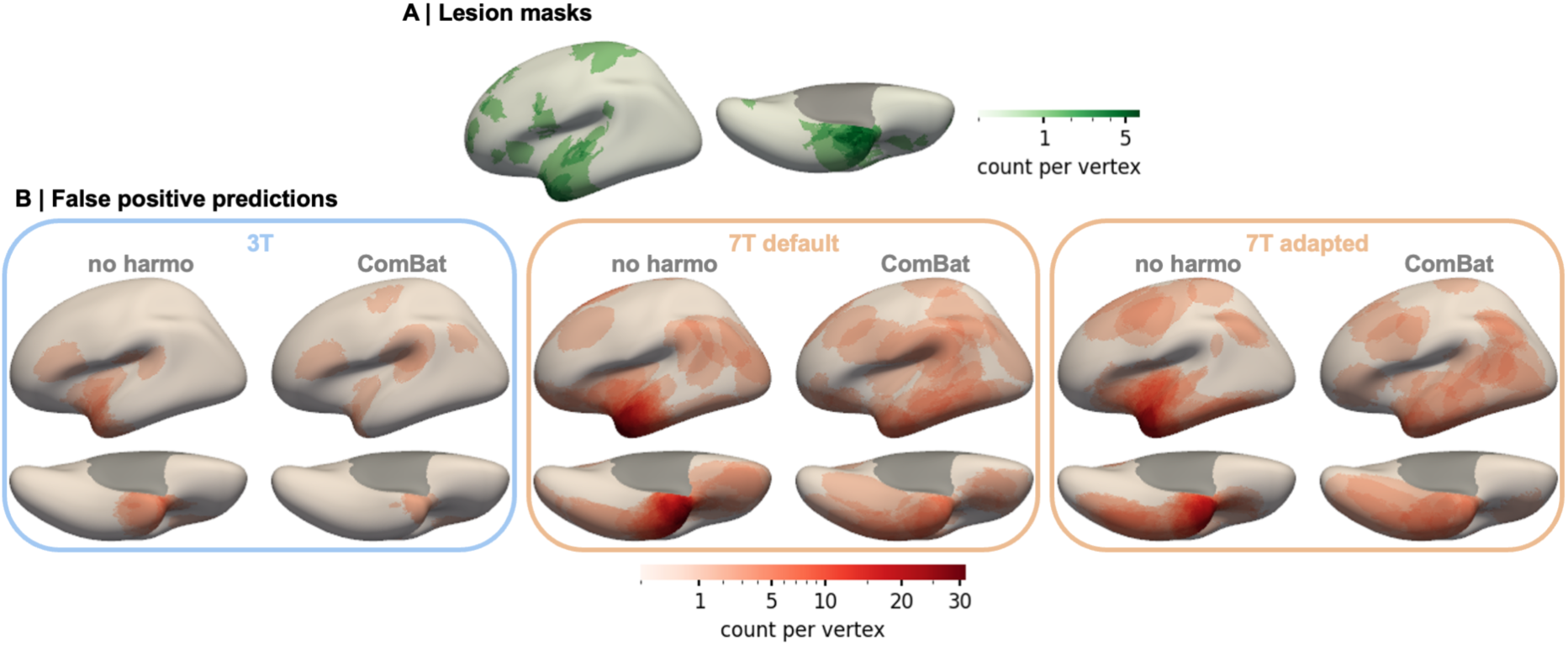
Spatial distribution of (A) manual FCD lesion masks and (B) false-positive MELD-graph predictions, based on 3T (left), 7T default (middle) and 7T adapted (right sub-panel) input/processing conditions. Masks or predictions were mapped and co-registered to a template left hemisphere cortical surface mesh, summed vertex-wise and shown in a power-law-scaled colourmap for visualization. Most FCDs in this dataset are located in the temporal and frontal lobes, a distribution that has also been described previously in larger datasets [Wagstyl et al., 2022]. False-positive predictions also predominantly occurred in the temporal lobes, especially at 7T and without ComBat harmonisation, so that reduced specificity or positive predictive value of predictions in this location should be taken into account when interpreting MELD-graph model outputs.

### Comparison of surface morphometry features

Mean normalised surface morphometry features in the manual FCD lesion labels were not significantly different across the field strength and processing conditions (3T, 7T default, 7T adapted; without and with ComBat harmonisation each) as shown in Supplementary Figure 5. Across healthy controls, differences in surface morphometry features between field strengths showed distinct spatial patterns (Figure 5, Supplementary Figure 6) that were largely compensated by ComBat harmonisation.

**Figure 5:**
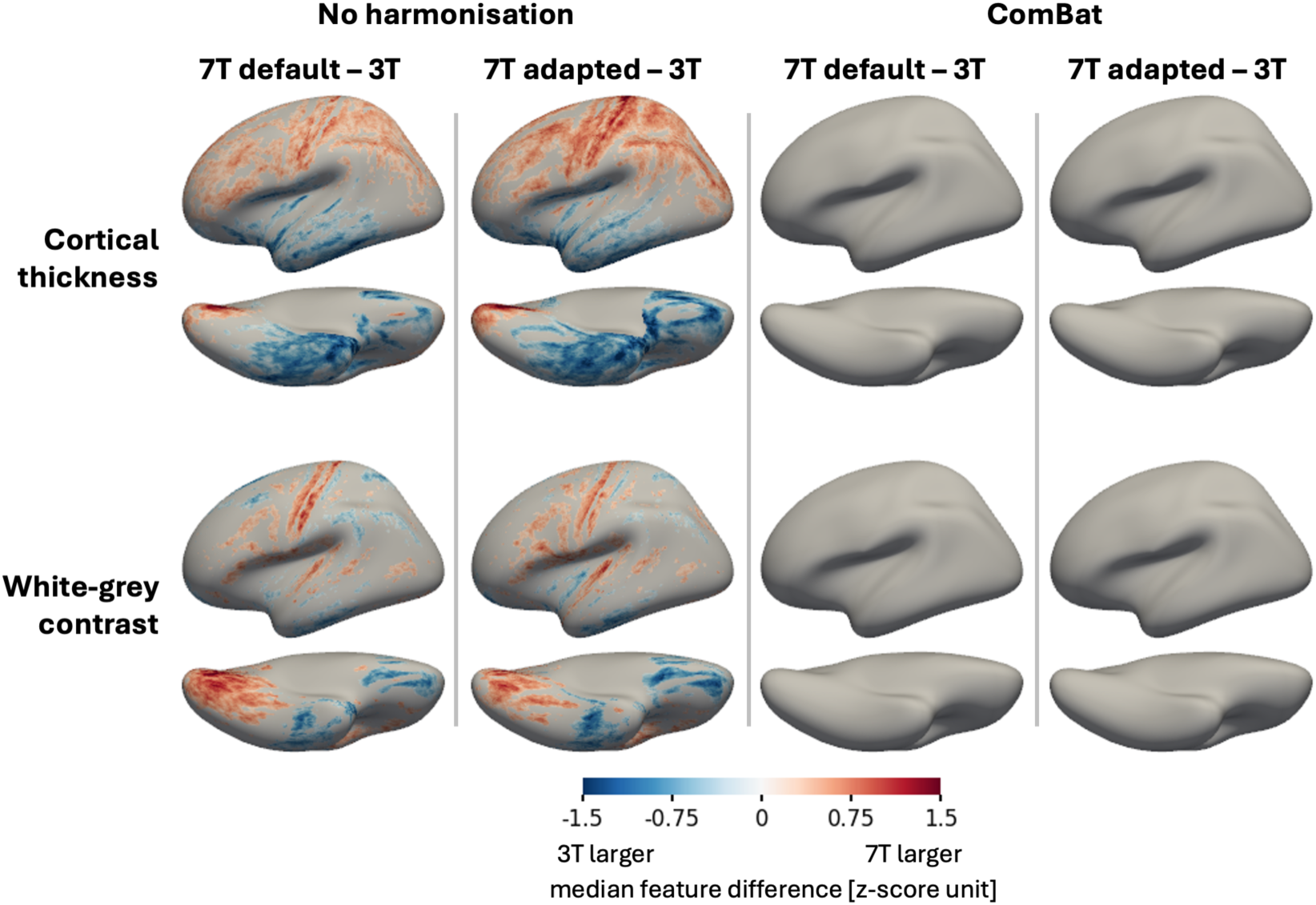
Spatial distribution of field-strength dependent differences in key surface morphometry features of MELD-graph. Across all healthy controls, per-vertex differences of cortical thickness (top row) and white-grey matter contrast (bottom row) between different conditions were computed: 7T default vs. 3T and 7T adapted vs. 3T reconstructions, each without or with ComBat harmonisation, respectively (columns). Features were mapped to the fsaverage_sym template, normalised by intra- and inter-subject z-scoring, as part of the MELD-graph pipeline, and additionally averaged across left and right hemisphere. For each vertex, a Wilcoxon signed rank test was computed; the median feature difference is plotted only for vertices with p_FDR_<0.05. Field-strength dependent differences are visible in the central and mesial occipital areas, possibly matching unimodal cortex and reflecting different cortical myelination, and also in the basal parts of the temporal and frontal lobe, where image inhomogeneity artefacts might be the underlying reason. Harmonisation compensates for field-strength specific biases. Supplementary Figure 6 shows plots for the other morphometry features (mean curvature, intrinsic curvature and sulcal depth).

## Discussion

We present a multi-centre validation study of the open-source research software MELD-graph for automated detection of FCD using 7T MRI compared to paired 3T MRI. We also investigate whether commonly proposed adaptations for processing ultra-high field MRI data using FreeSurfer, which is a key dependency for feature extraction in MELD-graph, can improve its performance. Images acquired at 3T and 7T, using both processing options, yielded comparable sensitivity for FCD detection of 52-65%. However, there were significantly more false-positive predictions per subject at 7T than at 3T (mean 0.6 vs. 0.2), as well as lower specificity at 7T compared to 3T (39%/45% vs. 76%). Most false-positives were located at the basal temporal lobes and temporal poles. ComBat harmonisation for site and scanner-specific effects improved specificity.

Although the small number of 3T-MRI negative cases does not allow statistically robust conclusions, it is notable that MELD-graph at 7T identifies an additional FCD case in this subgroup. Rather than automated lesion detection identifying only the visually most conspicuous cases, this would be consistent with the hypothesis that visual radiological review and automated analysis complement one another [Walger et al., 2024]. In clinical practice, 7T MRI is usually not performed in isolation but indicated as an additional investigation in 3T-MRI negative cases with a strong suspicion for an epileptogenic lesion or to characterise ambiguous 3T findings [Opheim et al., 2021], where it can not only identify new lesions, but also successfully disambiguate between true pathology and artefact [Klodowski et al., 2025]. We show performance metrics for a combination of 3T and 7T MELD-graph outputs (Supplementary Figure 2) with a slightly higher sensitivity and, as a trade-off, a higher number of false-positives, which increases the burden of manual review and reduces certainty.

While MELD-graph prediction extent and confidence varied between 7T and 3T in individual cases (Figure 3), we did not observe overall statistically significant differences in the size, surface-based dice scores, or prediction confidence scores (Supplementary Figure 1). It should be noted that the size of MELD-graph predictions has not been validated. It is likely affected by high interrater variability in manual segmentations of challenging lesions in both the training and test datasets, as well as design decisions that optimise for lesion detection rather than segmentation. This includes adaptive thresholding to reveal low-confidence predictions and an auxiliary distance loss that reduces the penalty for predictions adjacent to the lesion mask..

Surface-based brain morphometry using 7T MRI data is well known to face specific challenges and biases. These can be attributed to factors including smaller voxel sizes, artefacts in the basal frontal and temporal lobes, and the MP2RAGE sequence [Fujimoto et al., 2014; Lüsebrink et al., 2013]. Various adaptations have been proposed to improve Freesurfer segmentation accuracy at 7T, all implementing one or several of these common strategies: denoising of uniform MP2RAGE images, improved bias field correction, improved skullstripping and processing at sub-millimetric resolutions [Fujimoto et al., 2014; Lenge et al., 2026; Lüsebrink et al., 2013; Schwarzer et al., 2025]. Here, we evaluated the default Freesurfer pipeline as well as an adapted version with additional bias field correction, additional skullstripping and sub-millimetric segmentation. We show examples of reduced segmentation errors using the adapted pipeline (Figure 3, Supplementary Figure 4). Although there were no statistically significant differences in downstream MELD-graph performance for FCD detection, there was a trend favouring the adapted pipeline (Figure 2).

Very similar 7T-specific adaptations in the MELD-graph pipeline (bias field correction via SPM and FreeSurfer’s ‘-hires’ flag) were recently proposed and evaluated on a dataset including 87 DRFE patients [Lenge et al., 2026]. Overall, a sensitivity of 30/70 (43%) at 3T vs. 40/70 (57%) at 7T was reported, and more specifically, 25/37 (77%) in 3T MRI-positive patients and 6/12 (50%) in histopathologically verified FCD, while specificity in controls was 9/10 (90%). We additionally used SynthStrip for skullstripping, because we observed that this reduced mis-segmentations of the dura mater. However, our results are qualitatively different. We find lower specificity at 7T and overall no convincing evidence of increased detection performance using 7T data. This difference in performance may stem from different evaluation procedures between the studies.: Lenge et al., excluded predictions in manually identified regions of segmentation errors from the quantitative analysis. While this quality control step is well justified and likely improves accuracy, it also adds variability and workload in a clinical setting. Here, we measure performance without manual filtering, providing a conservative and unbiased estimate of specificity for MELD-graph.

Most false-positive findings at 7T were located in the basal temporal lobes (Figure 4B). Image inhomogeneity artefacts at 7T are likely underlying the observed spatial pattern. However, focal cortical dysplasias also frequently occur in the temporal lobes (Figure 4A) [Lamberink et al., 2020; Wagstyl et al., 2022]. The higher likelihood of temporal lobe false positives for MELD-graph predictions at both 3T and 7T should be taken into consideration for the clinical interpretation of the software’s outputs.

Although ComBat harmonisation compensated for 7T-specific biases in features (Figure 5), it did not recover specificity performance at 3T. We hypothesise that this is due to mis-segmentations in individual cases in different locations, for example of dura mater as cortex. This would not result in a systematic bias, but still cause false-positive predictions.

MELD-graph detection performance in our cohort when using 3T data – 65% sensitivity and 76% specificity, is similar to that reported in the original publication (70% sensitivity, 60% specificity in the main test set, and 72%/56% in an additional independent test cohort). Other independent studies using 3T MRI reported 76%/47% [Kaas et al., 2026], 63%/56% [Dell’Orco et al., 2026] or a sensitivity of 94% [Bücheler et al., 2026]. Differences in these performance figures can be attributed to underlying selection biases, different image acquisition protocols, as well as heterogeneous ground truth definitions used to evaluate predictions. For this reason, the present study was designed as a paired comparison between 3T and 7T specifically to assess generalisability.

Higher diagnostic performance of MELD-graph for cases with FLAIR available was reported in the original publication, though this was not a controlled experiment, and might be due to a bias in the selection of cases [Ripart et al., 2025]. It has been reported that quality of FLAIR sequences has an impact on detection performance of MELD-graph [Dell’Orco et al., 2026]. In this dataset – which includes heterogeneous acquisition protocols at 3T and 7T – we did not observe an increase in detection performance when using both FLAIR and T1-weighted (T1w) images as inputs, where available, compared to using T1w images only. On the contrary, more false-positive predictions resulted at 7T when using FLAIR (Supplementary Figure 3).

This study’s main limitation lies in the relatively small number of 23 FCD cases that could be included, which impacts the statistical power to detect smaller differences in detection performance. Furthermore, only 6 FCDs were histopathologically verified, whereas clinical diagnosis after radiologist review and multidisciplinary discussion served as ground truth in the remaining 17 cases. Radiologically 3T and 7T-MRI negative but histopathologically proven FCDs are valuable for assessing clinical utility of computer-aided lesion detection. However, these are also exceedingly rare. In the present cohort, one case was radiologically 3T and 7T MRI-negative with a histopathologically verified FCD. Data on histopathological subtypes in operated patients was not consistently available from the different sites, which precluded analyses stratified by this variable.

Future work is likely to lead to significant improvements in automated epileptogenic lesion detection at 7T. First, optimisation and standardisation of sequences at 7T [Opheim et al., 2021], alongside improvements to parallel transmit technology may further reduce image artefacts and improve inter-site consistency in image characteristics [Faber et al., 2026; Klodowski et al., 2025]. Second, improved brain segmentation and morphometry at 7T could be achieved either by generalist tools that are more robust to 7T-specific challenges while making full use of higher resolution images [Billot et al., 2023; Henschel et al., 2022] or dedicated software optimized for 7T data [Bazin et al., 2014; Gulban et al., 2018]. Finally, larger multi-centre 7T epilepsy datasets will be necessary to train and evaluate software for this specialised clinical application. The current MELD-graph adapted pipeline is available in the interim to inform the search for FCDs at 7T, and to provide a useful benchmark against which future developments can be compared.

In conclusion, an adapted version of MELD-graph that is available open-source can provide FCD lesion predictions using 7T MRI data at sub-millimetric resolution. We recommend using ComBat harmonisation for site-specific effects, and also evaluating predictions based on T1-weighted images only when FLAIR is available. 7T MELD-graph outputs may provide clinically useful information in challenging cases, though results should be interpreted with caution, critically re-evaluated by an expert human observer, and assessed by a multi-disciplinary team for concordance with seizure semiology, interictal and ictal EEG, and metabolic imaging (e.g. FDG-PET), given the lower specificity and higher number of false-positives especially in the temporal lobes.

## Data Availability

We are unable to make the MRI dataset publicly available due to data protection restrictions. The code used for analyses is openly available at the links (github) provided in the manuscript.

## Acknowledgements

CK is supported by the DFG and the Faculty of Medicine Tuebingen (MINT-CS), the Else Kroener Fresenius Stiftung (PRECISE.net), the Michael Foundation, de.NBI Cloud within the German Network for Bioinformatics Infrastructure (de.NBI) and ELIXIR-DE. CK, MR and KW are supported by the Wellcome Trust (301991/Z/23/Z). SA is supported by Vera Down Grant, BMA Foundation. MR and SA are supported by NIHR (503216). RJP is supported by GOSH Children’s Charity. TB is supported by GOSH Biomedical Research Centre (BRC). JSD is supported by NIHR. MHE is supported by The Foulkes Foundation. JOC is supported by the Royal College of Radiologists. TB is supported by Neuro-aCSis Bonn Neuroscience Clinician Scientist Programme (German Research Foundation, Deutsche Forschungsgemeinschaft, DFG, 493623632). TR is supported by the German Ministry for Research, Technology and Space (epi-center.ai). KK, CTR, TEC are supported by the Medical Research Council: Developmental pathway funding scheme UKRI790, Mid-range equipment fund award UKRI2679, University of Cambridge Impact Acceleration Account Award MR/X502844/1, Clinical Research Infrastructure Award MR/M008983/1, the Cambridge University Hospitals Academic Fund for functional neurosurgery, the NIHR Cambridge Biomedical Research Centre (NIHR203312). This article represents independent research in part funded by the infrastructure of the National Institute for Health Research (NIHR) Mental Health Biomedical Research Centre (BRC) at South London and Maudsley NHS Foundation Trust and King’s College London. This research was funded/supported by the National Institute for Health and Care Research (NIHR) Clinical Research Facility at Guy’s and St Thomas’ NHS Foundation Trust. The content of this manuscript is solely the responsibility of the authors, the views expressed are those of the authors and not necessarily those of the NHS, the NIHR, the Department of Health and Social Care or any of the other funders.The funders had no role in study design, data collection and analysis, decision to publish or preparation of the manuscript.

## Author contributions

*Cornelius Kronlage:* Conceptualization, Methodology, Investigation, Writing - Original Draft. *Mathilde Ripart:* Methodology, Writing - Review & Editing. *Rory J Piper:* Data Curation, Writing - Review & Editing. *Chiara Casella:* Data Curation, Writing - Review & Editing. *Krzysztof Klodowski:* Data Curation, Writing - Review & Editing. *Tobias Bauer:* Data Curation, Writing - Review & Editing. *Maria H Eriksson:* Data Curation, Writing - Review & Editing. *Philippa Bridgen:* Data Curation, Writing - Review & Editing. *Martin M Tisdall:* Data Curation, Writing - Review & Editing. *Simon R Bouschery:* Data Curation, Writing - Review & Editing. *Annalena Lange:* Data Curation, Writing - Review & Editing. *Eberhard D Pracht:* Data Curation, Writing - Review & Editing. *Tony Stöcker:* Data Curation, Writing - Review & Editing. *Jon O Cleary:* Data Curation, Writing - Review & Editing. *Felice D’Arco:* Data Curation, Writing - Review & Editing. *Torsten Baldeweg:* Resources, Writing - Review & Editing. *John S Duncan:* Resources, Writing - Review & Editing. *Rainer Surges:* Resources, Writing - Review & Editing. *Theodor Rüber:* Resources, Writing - Review & Editing. *Christopher T Rodgers:* Resources, Writing - Review & Editing. *Thomas E Cope:* Resources, Writing - Review & Editing. *David W Carmichael:* Resources, Writing - Review & Editing. *Jonathan O’Muircheartaigh:* Data Curation, Resources, Writing - Review & Editing. *Sophie Adler:* Conceptualization, Methodology, Resources, Project administration, Writing - Review & Editing. *Konrad Wagstyl:* Conceptualization, Methodology, Resources, Project administration, Writing - Review & Editing.

