## Supplementary Material for "Multi-centre validation of MELD-graph for automated focal cortical dysplasia detection using 7T compared to paired 3T MRI"

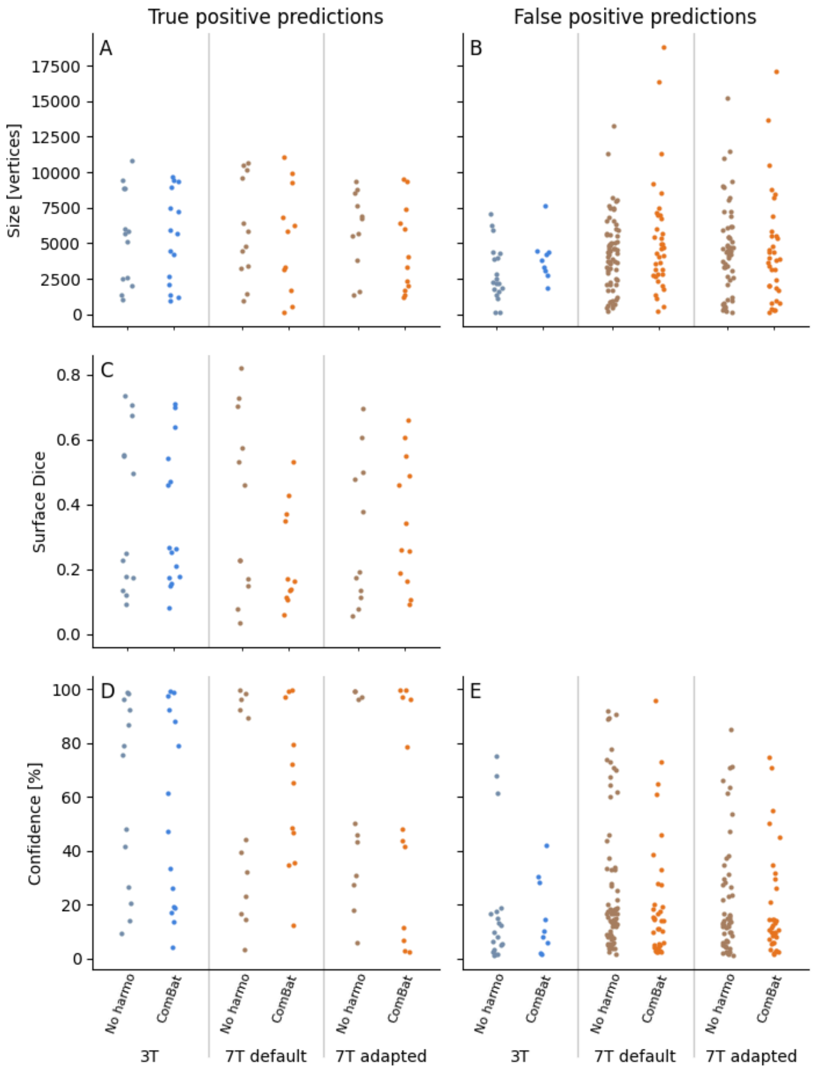


Supplementary Figure 1: Characteristics of MELD-graph predictions at 7T vs. 3T. (A, B) Size (measured by the number of cluster vertices), (C) Dice score between ground truth (on the template surface) and MELD-graph prediction, and (D, E) confidence score (in %) of true-positive (A, C, D) and false-positive (B, E) MELD-graph predictions, when using 3T vs. 7T T1-weighted images as inputs and default or adapted processing, without and with ComBat harmonisation. The distributions in each case are not statistically significantly different (Kruskal-Wallis test $p > 0.05$).

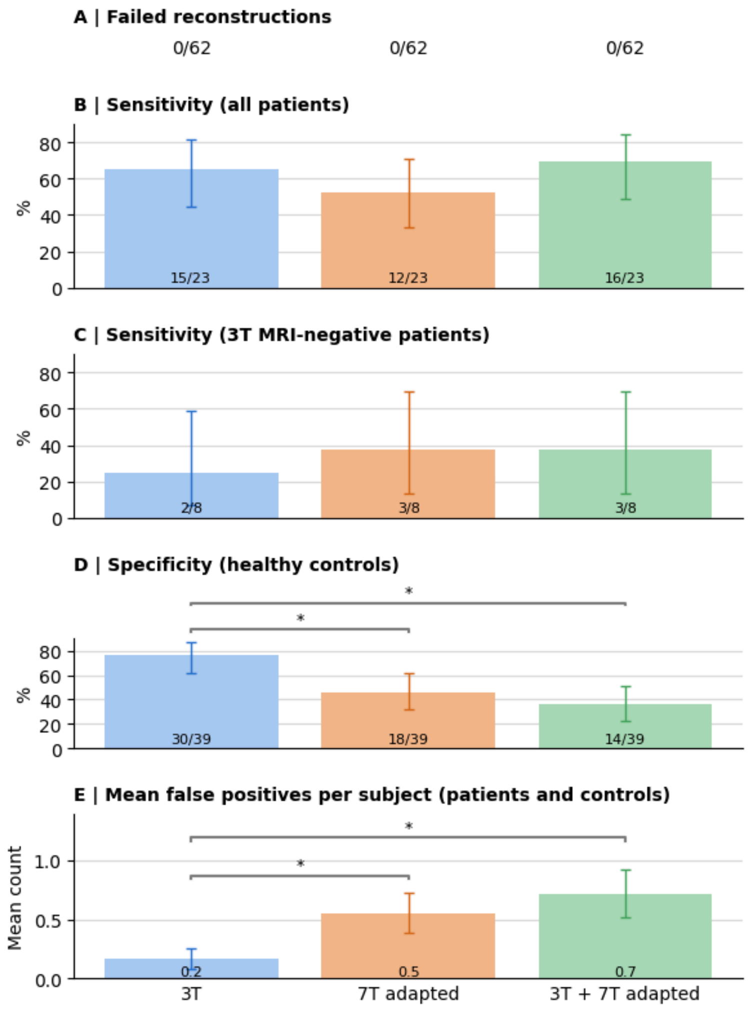


Supplementary Figure 2: FCD detection performance using combined 3T and 7T MELD-graph predictions. Bar plots show performance observed using ComBat harmonisation and T1-weighted images acquired at 3T (left), 7T with adapted preprocessing (middle) and a combination of 3T and 7T findings (right). For the combination, an FCD was considered ‘detected’ if there was an overlapping prediction at either field strength; false-positive predictions were summed. (A) Fraction of failed Freesurfer reconstructions in each condition; further metrics below are only for n=61 subjects with successful reconstructions. (B) Sensitivity in all FCD patients. (C) Sensitivity in radiologically 3T MRI-negative patients. (D) Specificity. (E) Mean number of false positive predictions in patients and controls. Error bars show 95% confidence intervals. *statistically significant differences ($p < 0.05$ with Benjamini-Hochberg false discovery rate correction)

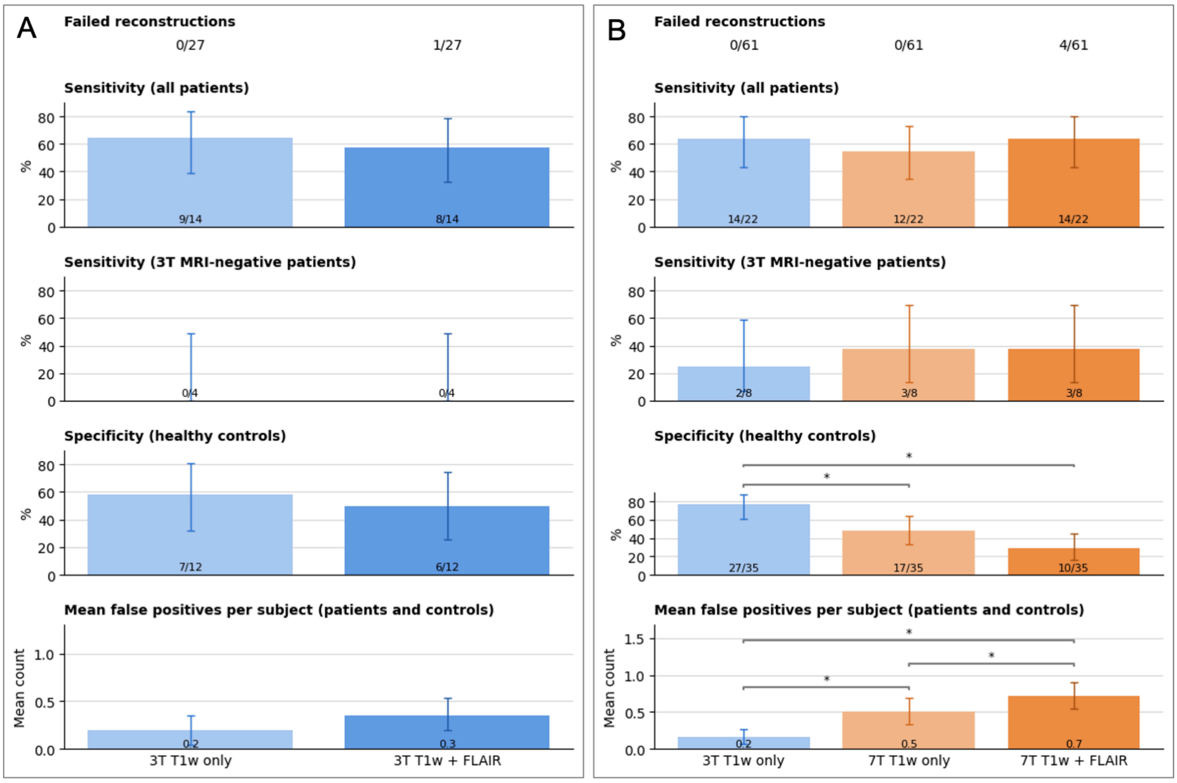


Supplementary Figure 3: FCD detection performance of MELD-graph using only T1-weighted images compared with FLAIR and T1-weighted (T1w) images. (A) Subset of n=27 subjects with 3T FLAIR images available: T1w only (left), T1w and FLAIR (right). (B) Subset of n=61 subjects with 7T FLAIR available: 3T T1w only (left), 7T T1w only (middle), 7T T1w and FLAIR (right), in each case with ComBat harmonisation. Error bars show 95% confidence intervals. *statistically significant differences ($p < 0.05$ with Benjamini-Hochberg false discovery rate correction). There is a trend for the addition of FLAIR to result in lower specificity. At 7T, there are significantly more false-positive predictions when using FLAIR compared to using T1w only.

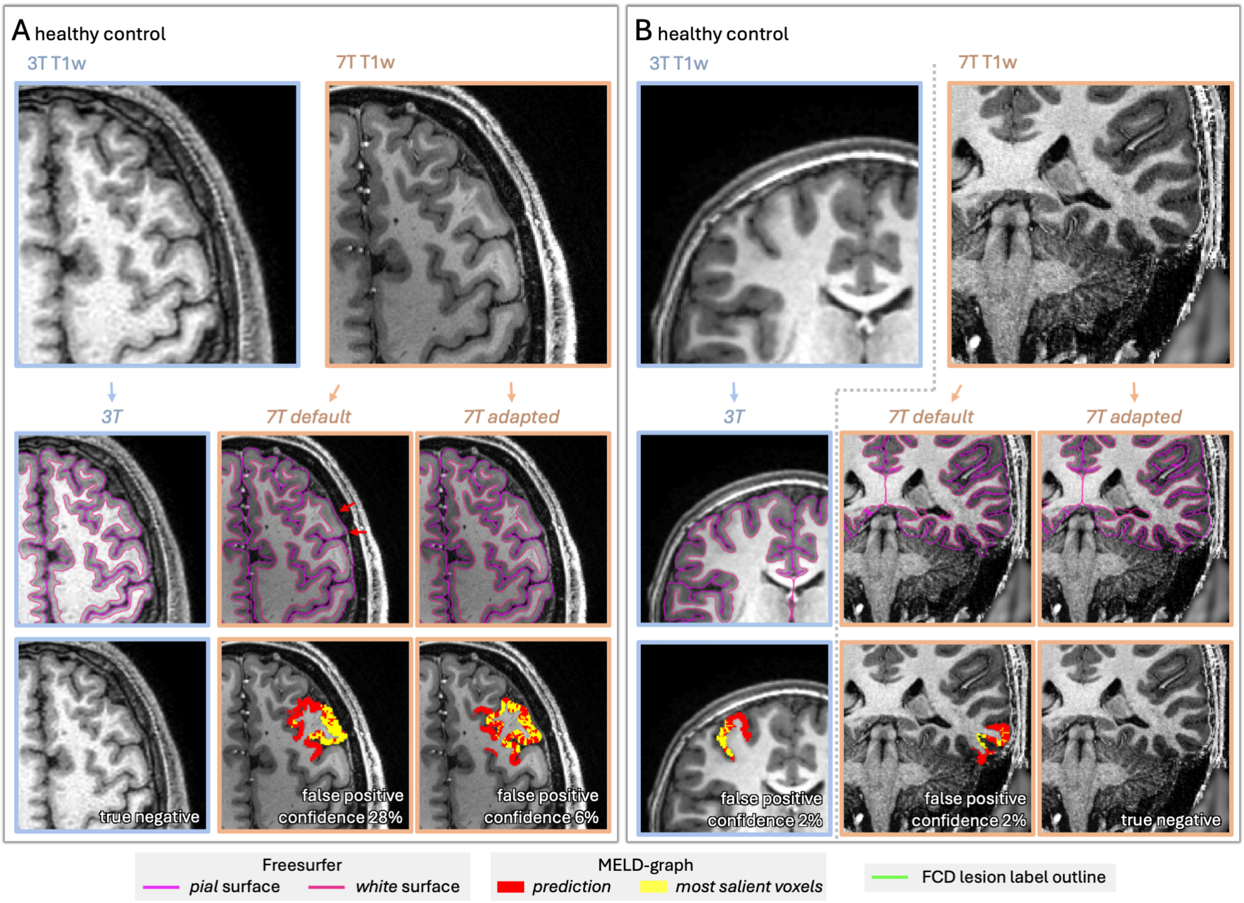


Supplementary Figure 4: Further case examples of false-positive MELD-graph predictions, all obtained using ComBat harmonisation. (A) Healthy control age 21-25 years, with false-positive predictions at 7T but not 3T. (B) Healthy control age 6-10 years, with false-positive predictions in different locations for 3T and 7T default, but not 7T adapted. For each case, input images without overlays, with overlaid lesion label outlines (green outline) and MELD-graph predictions (red/yellow clusters), and input images with overlaid Freesurfer surfaces and red arrows showing segmentation errors are displayed.

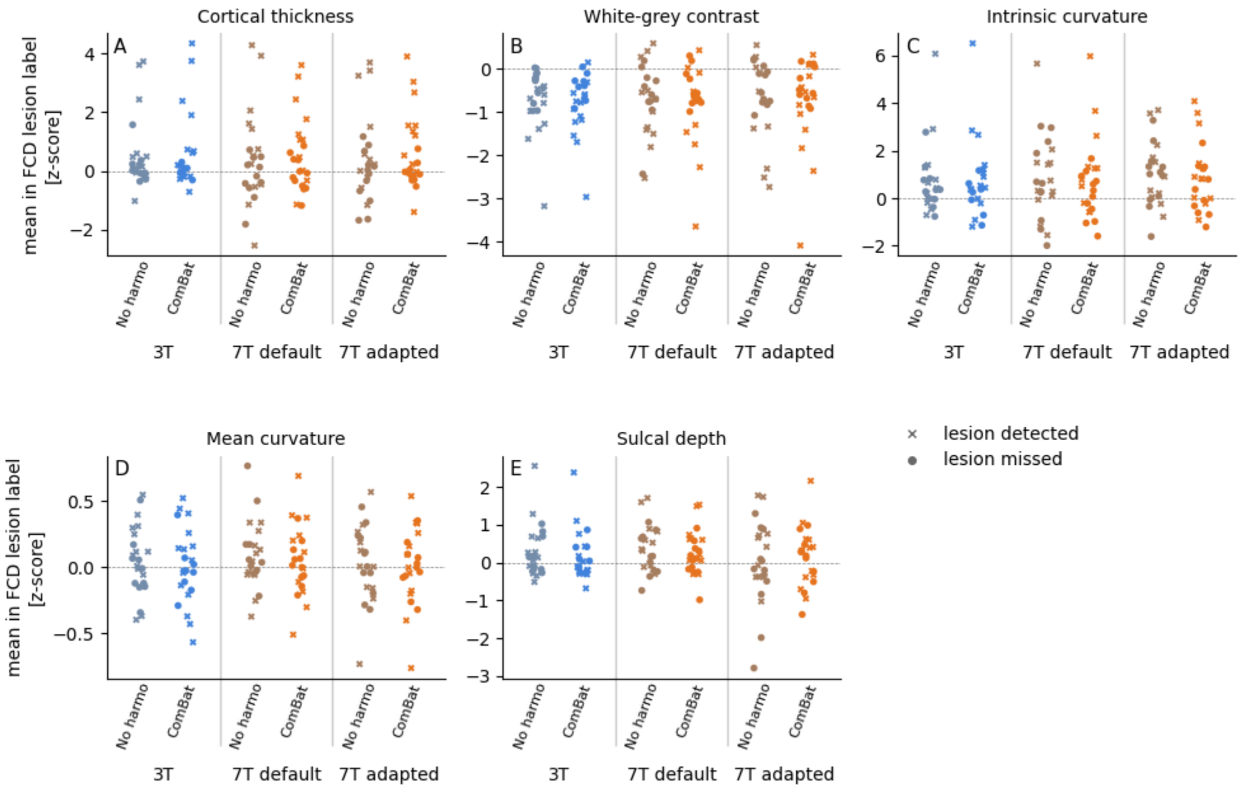


Supplementary Figure 5: Normalised surface morphometry features (mean curvature, intrinsic curvature, sulcal depth, cortical thickness, white-grey contrast) in the FCD lesions (mean per label) for ‘3T’, ‘7T default’ and ‘7T adapted’ input/processing conditions, each without and with ComBat harmonisation. The shape of each point (cross/circle) indicates whether the lesion was detected by MELD-graph in this condition. There were no statistically significant differences in distributions between all ‘No harmo’ and all ‘ComBat’ conditions, respectively (Friedman-test $p > 0.05$).

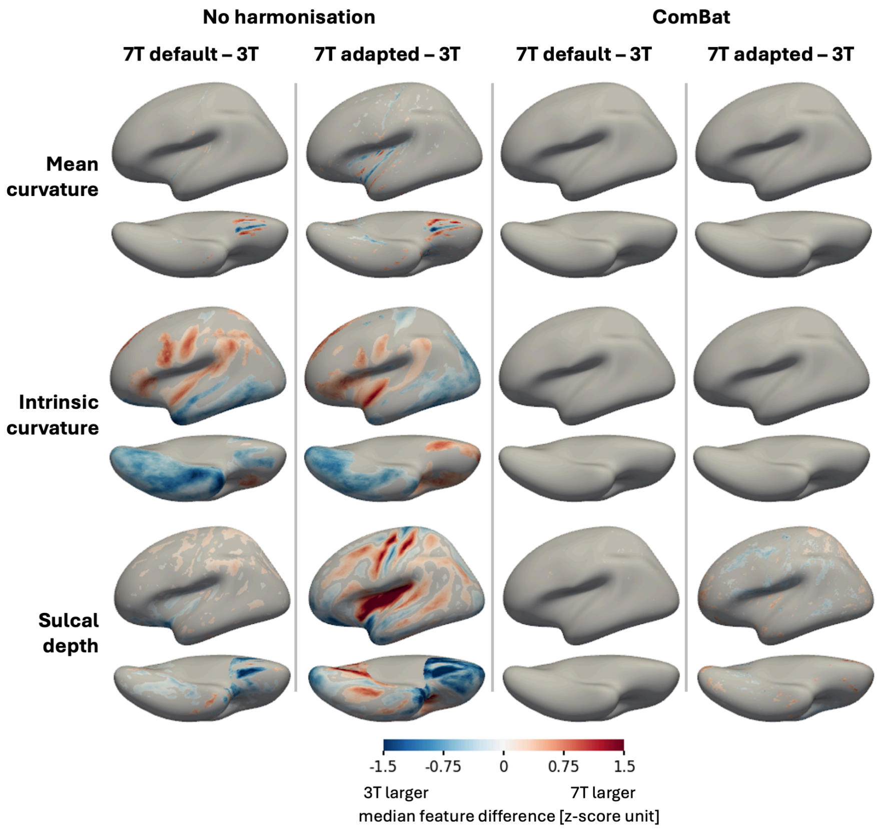


Supplementary Figure 6: Spatial distribution of field-strength dependent differences in additional surface morphometry features of MELD-graph. Across all healthy controls, per-vertex differences of mean curvature (top row), intrinsic curvature (middle row) and sulcal depth (bottom row) between different conditions were computed: 7T default vs. 3T and 7T adapted vs. 3T reconstructions, each without or with ComBat harmonisation, respectively (columns). Features were mapped to the fsaverage_sym template, normalised by intra- and inter-subject z-scoring, as part of the MELD-graph pipeline, and additionally averaged across left and right hemisphere. For each vertex, a Wilcoxon signed rank test was computed; the median feature difference is plotted only for vertices with $p_{FDR}<0.05$. Harmonisation compensates for field-strength specific biases.
